# Long-term Exposure to Air Pollution and Lipometabolic Disturbance: A Systematic Review and Meta-Analysis

**DOI:** 10.1101/2020.05.19.20106849

**Authors:** Haohao Chen, Ye Zhu, Liuhua Shi, Andrew Rosenberg, Lixin Tao, Xingfan Zhou, Rui Chen, Ji Wang

## Abstract

**Objectives:** To reveal the chronic effects of air pollutants on lipometabolic disturbance via detectable lipoprotein parameters.

**Study Design:** This is a systematic review and meta-analysis.

**Methods:** Seven online databases were searched to conduct a meta-analysis of epidemiological studies examining the relationship between air pollution and lipid parameter levels. Subgroup analysis was additionally carried out for each air pollutant studied.

**Results:** A total of 2,274 records were retrieved, resulting in 10 studies included in the final quantitative meta-analysis, comprising seven studies in Europe and the United States and three studies in mainland China. Using a random-effect model, the results showed that for each 10 μg/m^3^ increment in PM_2.5_, TC, LDL-C, and HDL-C levels and metabolic syndrome (MetS) incidence increased by 3.31% (95% CI: 2.29%, 8.91%), 2.34% (95% CI: 1.30%, 3.39%),-1.57% (95% CI: −1.85%, −1.28%), and 4.33% (95% CI: 2.69%, 5.98%), respectively; for each 10 μg/m^3^ increment in PM_10_, LDL-C, TG, HDL-C levels increased by 5.27% (95% CI: 2.03%, 8.50%), −0.24% (95% CI: −0.95%, −0.47%), and 0.45% (95% CI: −0.57%, 1.47%), respectively; for each 10 μg/m^3^ increment in NO_2_, TG and HDL-C levels increased by 4.18% (95% CI: 1.12%, 7.23%) and −0.51% (95% CI: −2.61%, 1.58%), respectively. No significant associations were detected for combinations of air pollutants on lipometabolic disturbance.

**Conclusion:** Increased air pollutant exposure is significantly associated with fluctuation in blood lipid parameter levels, which can be an indicator of the body’s lipometabolic disturbance.

## 1. Introduction

Air pollution is among the greatest environmental risks to global health^1-3^. The World Health Organization (WHO) estimates that ambient air pollution is responsible for nearly seven million premature deaths worldwide every year^4^. Major components of atmospheric pollution include particulate matter of different diameter sizes, such as PM_2.5_, PM_10_, UFP (ultrafine particulate matter), and gas compounds such as CO, NO_x_, SO_2_, O_3_, among others. Primarily deriving from fuel combustion, industrial emissions, transportation and ground dust, these pollutants can cause serious harm to the human body^5^. A growing body of evidence demonstrated the links between air pollution and cardiovascular diseases^6-9^, respiratory diseases^10-13^, and neurological disorders^14-16^, among which cardiovascular disease is the leading cause of death in low-income and middle-income countries^17, 18^. Conventionally, hazardous factors for cardiovascular diseases (CVD) have been partly connected with abnormal levels in lipoprotein-lipid parameters, including high density lipoprotein cholesterol (HDLC), low density lipoprotein cholesterol (LDL-C), total cholesterol (TC), triglycerides (TG), total cholesterol (TC) and so on, which may indicate lipometabolic disturbance^19, 20^. The co-occurrence of these changing lipid parameters, known as one risk of metabolic syndrome (MetS), may therefore amplify susceptibility to the CVD risks associated with air pollution exposure^21-24^.

In recent years, mounting studies have researched the associations between air pollution and lipid profile parameters, adding to evidences that suggests air pollution may contribute to changes in blood lipid parameters^25-29^. However, most previous studies have been limited to single air pollutants. To date, only one study^25^ has been found which offers a synthesis of the links between atmospheric pollutants and lipid parameters, but it only included 3 studies for the quantitative meta-analysis. The aim of our meta-analysis is to provide a more comprehensive and quantitative overview of the literature regarding of the association between various air pollutants, lipid parameters, and MetS morbidity.

## 2. Materials and Methods

### 2.1 Search Strategy

We conducted the study using standard methods deriving from the Preferred Reporting Items for Systematic Review and Meta-Analysis (PISMA)^30, 31^. Seven electronic databases, including China National Knowledge Infrastructure (CNKI), Wanfang, Vip, SinoMed, Pubmed, EMBASE, and the Cochrane Library, were searched for peer-reviewed articles published from 2000 to March 28, 2020. Keywords searched included: (“Air Pollution” OR “Pollution, Air” OR “Air Quality” OR “Ultrafine Fibers” OR “Airborne Particulate Matter” OR “Particulate Matter, Airborne” OR “Air Pollutants, Particulate” OR “Particulate Air Pollutants” OR “Ambient Particulate Matter” OR “Particulate Matter, Ambient” OR “Ultrafine Particulate Matter” OR “Particulate Matter, Ultrafine” OR “Ultrafine Particles” OR “Particles, Ultrafine” OR “PM_2.5_” OR “PM_10_” OR “Nitrogen oxides” OR “NO_2_” OR “Sulphur dioxide” OR “SO_2_” OR “Carbon monoxide” OR “CO” OR “Black carbon”) AND (“Cardiovascular Disease” OR “Disease, Cardiovascular” OR “Diseases, Cardiovascular” OR “Metabolic syndrome” OR “MetS” OR “High density lipoprotein” OR “HDL” OR “Low density lipoprotein” OR “LDL” OR “cholesterol” OR “ cholesterin” OR “cholesteric” OR “TC” OR “TG” OR “dyslipidemia” OR “Hypertriglyceridemia” OR “Triglycerides” OR “hypercholesterol*”, “hypertriglycerid*” OR “HDL-C” OR “LDL-C”) AND (“Cohort” OR “Cross-sectional” OR “Case control” OR “Case-control” OR “Epidemiology OR “Epidemiological”). Additionally, references of included literature and one previously published systematic review were manually retrieved.

### 2.2 Eligibility Criteria

#### 2.2.1 Inclusion Criteria

Studies were included if: (1) The study includes all population with no age, gender, or race restrictions. (2) The study describes the relationship between air pollutants and metabolic disease/cardiovascular disease. (3) Results quantify changes in lipid parameters, including LDL-C, HDL-C, TC, TG, etc. (4) Cross-sectional studies, cohort studies, or case control studies. (5) The Literature is published in either Chinese or English.

#### 2.2.2 Exclusion Criteria

Studies were excluded if: (1) The research was based on animal experiments or cytological experiments. (2) The study was a meta-analysis or systematic review. (3) The study was a duplicate. (4) Effect estimates were not able to be conversed as percent changes in lipid parameters per 10 μg/m^3^ change in air pollutants. (5) The collected data resulted from an independent study. If inconsistent results have been reported for a given population or period at the time publications were retrieved, older studies were superseded by the most recent publication.

### 2.3 Literature Screening and Quality Evaluation

Two researchers independently retrieved and screened the literature following the above order and criteria. Upon review, a third researcher was consulted when discrepancies occurred. The methodological quality of each cohort study was evaluated in line with the Newcastle-Ottawa Scale (NOS) literature quality evaluation tool^32^, which includes three aspects of population selection, comparability, and results. A total of 8 items belonging to NOS could be graded by an “*” for each item, and the top score can add up to 9 “*”. Cross-sectional studies were assessed by the American Health Care Quality and Research Institute (AHRQ) scale for quality evaluation^33^. An 11-point scale is used to calculate responses of either “Yes”, “No”, or “Not clear”, for a total score of 11. Specifically for quality evaluation in this study, a study was awarded one “*”, corresponding to one point, for each NOS item it contained. “yes” was recorded as one point, whereas “no” or “uncertain” was recorded as zero for each component of the AHRQ scale.

### 2.4 Data extraction and statistical analysis

Information was independently extracted by two researchers based on a pre-set data extraction form which included: author name(s), year of publication, study period, location, population, median age, sample size, experimental design type, exposure time, and contaminants. Divergent interpretations were reconciled by a third researcher.

When the literature data were extracted, the percent changes in lipid parameters per 10 μg/m^3^ change in air pollutants, from original data or conversed data, were directly utilized. As such, the results presented in this meta-analysis are expressed as the percentage change in lipid parameters and MetS incidence for each 10 μg/m^3^ increment in air pollutant. The between-study heterogeneity was conducted on the basis of both the I^2^ statistic and Q-test^34^. High heterogeneity was considered if a study was evaluated as I^2^ ≥ 50% or the p-value for a Q-test was < 0.01, and pooling effect estimates was conducted by Der Simonian and Laird random effects model. On the contrary, moderate or low heterogeneity was considered if a study was evaluated as I^2^ < 50% or the p-value for a Q-test was ≥ 0.01, and pooling effect estimates was conducted by Mantel-Haenszel fixed effects model. Stata software version 14.0 was used in this study. A p-value < 0.05 was considered statistically significant.

## 3. Results

### 3.1 Literature Retrieval and Characteristics

A total of 2,274 records were obtained from multiple databases. After duplicates were removed, 1,174 records were screened by checking the titles and abstracts. 1,154 full-text publications were subsequently assessed using the eligibility criteria. As a result, ten records met all criteria and were pooled in the ultimate quantitative meta-analysis^26-29, 35-40^. The detailed study selection process is shown in Fig. 1.

**Fig.1.**
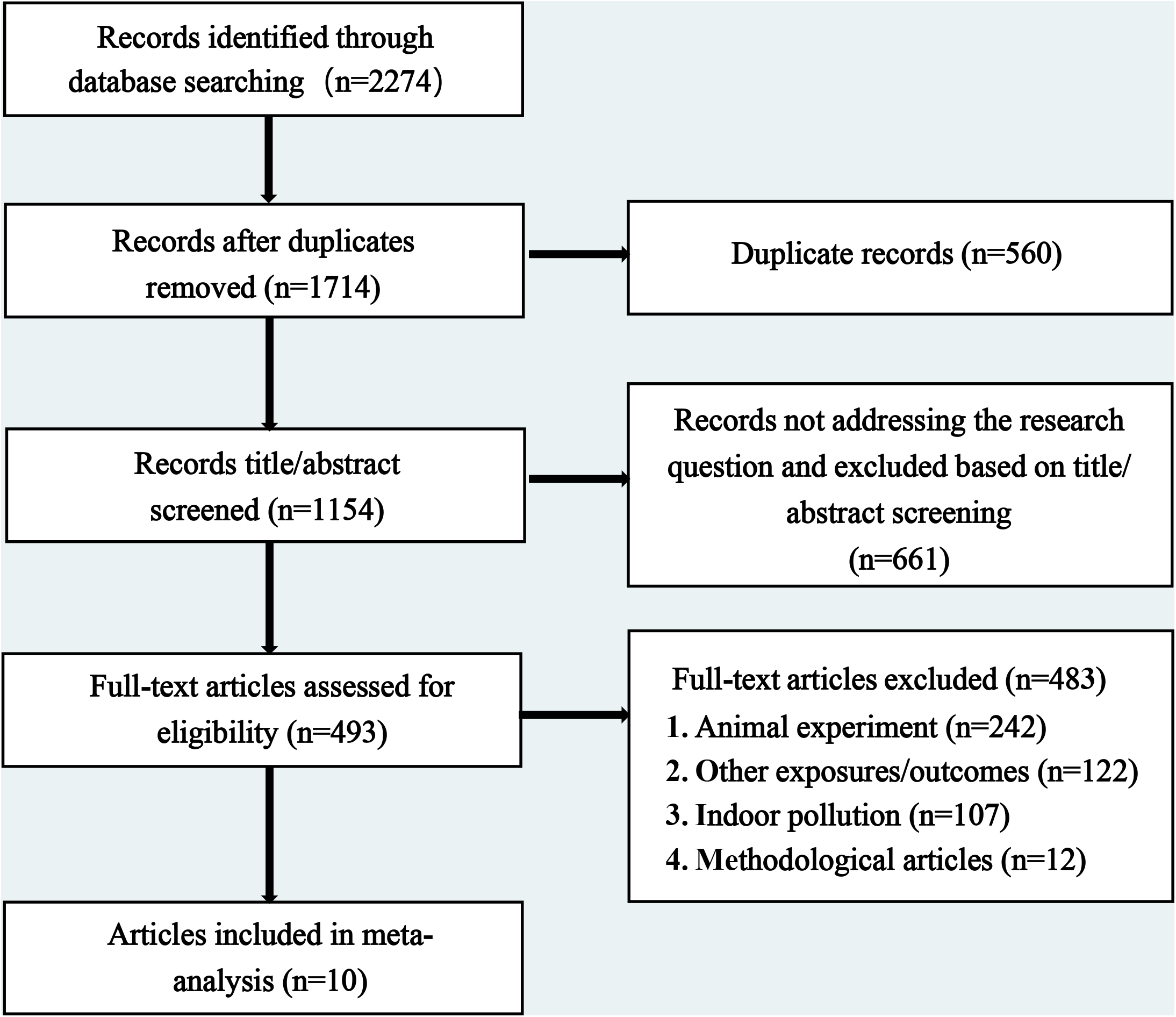
Flow diagram of study selection process.

Overall, this study includes a total of ten records comprising eight cohort studies^26, 27, 29, 35-38, 40^ and two cross-sectional studies^28, 39^. An exposure time of less than or equal to thirty days was recorded as short-term exposure, whereas an exposure time greater than thirty days was recorded as long-term exposure. Study characteristics and descriptions are summarized in Table 1~2.

**Table 1.** General characteristics of ten included studies.

| Author | Published year | Study period | Location | Study population | Sample size | Study design | Exposure duration | Follow-up | NOS score | AHRQ score |
| --- | --- | --- | --- | --- | --- | --- | --- | --- | --- | --- |
| Yang et al. | 2018 | 2006~2008 | Chian, Liaoning province | Adults, 18~74y | 15477 | cohort | Long-term | 3y | 8 |  |
| Lee et al. | 2019 | 2002~2013 | Korea, Seoul | Adults | 119998 | cohort | Long-term | 4y | 6 |  |
| Rachel et al. | 2016 | 1993~2011 | USA, New York | Adults, 70y | 587 | cohort | Long-term | 10y | 8 |  |
| Wang et al. | 2018 | 2011~2015 | China, Ganshu province | Adults, 60y | 3912 | cohort | Long-term | 4y | 7 |  |
| Wu et al. | 2018 | 1999~2005 | USA | Adults, 42~52y | 2289 | cohort | Long-term | 7y | 7 |  |
| Maayan et al. | 2016 | 2003~2012 | Israel | Adults, 60y | 73117 | cohort | Long-term | 10y | 7 |  |
| Hou et al. | 2020 | 2015~2017 | China, Henan province | Adults, 18~79y | 39089 | cross-sectional | Long-term | 2y |  | 10 |
| Shanley et al. | 2016 | 1988~1994 | USA | Adults, 41y | 11623 | cohort | Long-term | 3y | 8 |  |
| Cai et al. | 2017 | 2006~2013 | Norseland | Adults, >20y | 144082 | cohort | Long-term | 3y | 9 |  |
| Mette et al. | 2015 | 1993~1997 | Denmark, Copenhagen | Adults, 50~64y | 39863 | cross-sectional | Long-term | 5y |  | 9 |

Abbreviation: y indicates years

**Table 2.**
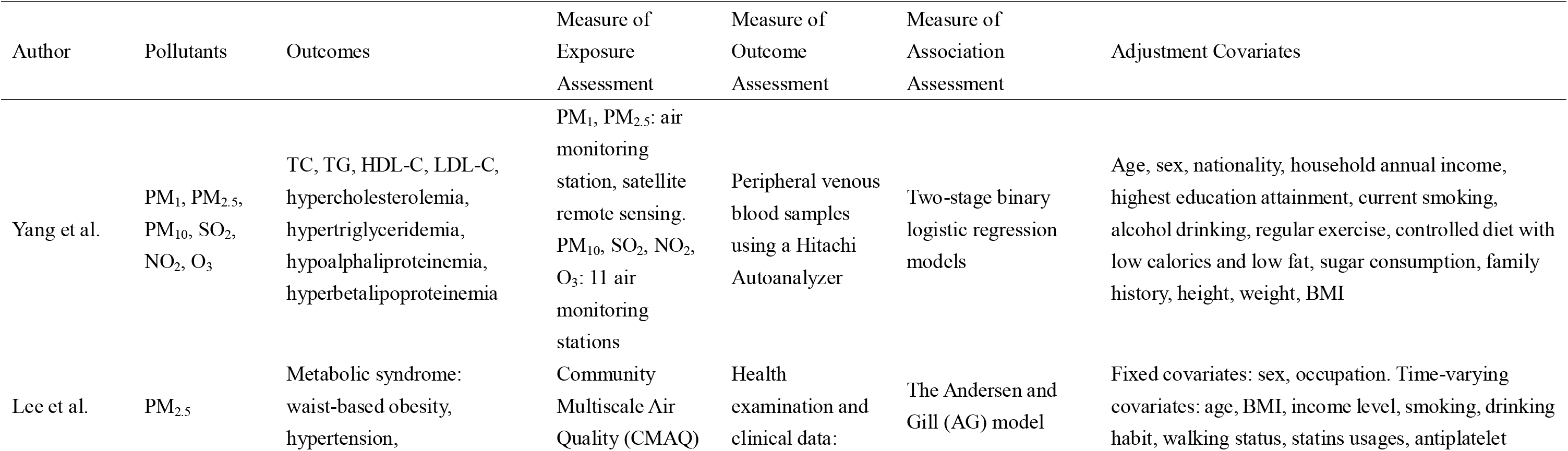
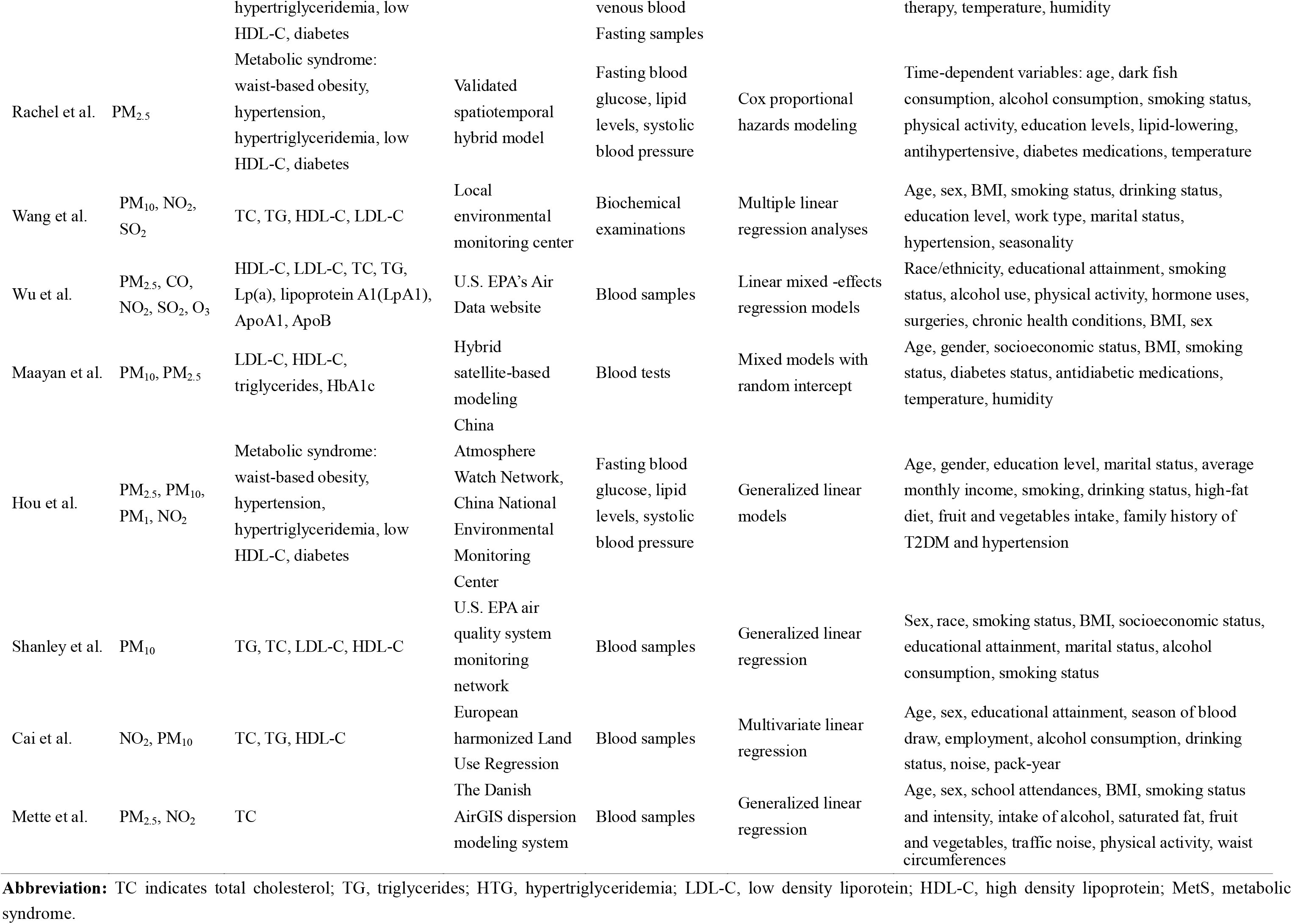
Description of pollutants, outcomes, measures, and adjustment covariates in included studies.

### 3.2 Quality Evaluation

Using the NOS scale to evaluate eight cohort studies, one study was less representative, three studies were less comparable, six mentioned follow-ups, eight mentioned the specific measurement methods from which the results were derived, and six were subject to the integrity of follow-up. Using the AHRQ scale to evaluate two cross-sectional studies, both mentioned the source of data, patient identification time period, follow-up situation, and continuity of the subject. Final scores based on the NOS scale and the AHRQ scale for each respective study are summarized in Table 1.

### 3.3 Data Extraction and Conversion

The results of the data extraction of the ten records are shown in Table 3. The change of air pollutant concentrations varied from study to study. As such, in order to pool data for further analysis, original data from each study was uniformly converted to the rate of change of each lipid parameter per 10 μg/m^3^ increment change in air pollutant. A complete list of converted results is reported in Table 4.

**Table 3.** Data extractions from included 10 studies.

| Author | Pollutant/<br>incremnet<br>($\mu\text{g}/\text{m}^3$) | % change in outcome (95% CI) per pollutant increment and morbidity in MetS | | | | | |
| --- | --- | --- | --- | --- | --- | --- | --- |
|  |  | TC | TG | HTG | LDL-C | HDL-C | MetS |
| Yang et al. | PM<sub>2.5</sub>/10 | 1.1(0.8, 1.4) * | 1.1(0.4, 1.8) * | 1.07(0.95, 1.19) | 2.9(2.4, 3.5) * | -1.1(-1.4, -0.8) * | / |
|  | PM<sub>10</sub>/ 10 | -0.2(-0.5, 0.1) | 4.7(3.6, 5.9) * | 1.14(1.01, 1.29)* | -0.9(-1.3, -0.4) * | -0.2(-0.7, 0.2) | / |
|  | NO<sub>2</sub>/ 7.4 | 0.7(0.0, 1.4) * | 6.0(3.5, 8.6) * | 1.21(0.76, 1.90) | -0.1(-0.7, 0.5) | -1.6(-2.3, -1.0) * | / |
| Lee et al. | PM<sub>2.5</sub>/10 | / | / | 1.47(1.42, 1.51) | / | -1.63(-1.69, -1.56) * | 1.07(1.03, 1.11) * |
| Rachel et al. | PM<sub>2.5</sub>/1 | / | / | 1.14(1.00, 1.32) * | / | -0.98(-1.13, -0.85) | 1.27(1.06, 1.52) * |
| Wang et al. | PM<sub>10</sub>/10 | 0.45(0.08, 0.82) * | -0.01(-1.01, 1.00) | / | 0.83(0.21, 1.45) * | 0.29(0.10, 0.49) * | / |
|  | NO<sub>2</sub>/10 | 1.16(-1.06, 3.43) | 5.58(-0.62, 12.16) | / | 39.01(31.43, 47.03) * | -3.55(-6.40, -0.61) * | / |
| Wu et al. | PM<sub>2.5</sub>/3 | 0.5(-0.1, 1.1) | / | / | 1.1(-0.2, 2.3) | -0.7(-1.4, -0.1) * | / |
| Maayan et al. | PM<sub>2.5</sub>/7<br>(normal) | / | / | 0.28(-0.03, 0.59) | 1.28(1.03, 1.52) * | -1.29(-1.43, -1.15) * | / |
|  | PM<sub>2.5</sub>/7<br>(diabetes) | / | / | 0.41(0.09, 0.74) * | 1.54(1.26, 1.83) * | -1.31(-1.47, -1.16) * | / |
|  | PM<sub>10</sub>/20<br>(normal) | / | / | 0.16(-0.12, 0.45) | 2.28(2.05, 2.50) * | -1.13(-1.26, -0.99) * | / |
|  | PM<sub>10</sub>/20<br>(diabetes) | / | / | 0.31(0.02, 0.61) | 2.37(2.11, 2.63) * | -1.13(-1.27, -0.99) * | / |
| Hou et al. | PM<sub>2.5</sub>/5 | / | / | / | / | / | 1.42(1.36, 1.49) * |
|  | PM<sub>10</sub>/5 | / | / | / | / | / | 1.23(1.20, 1.25) * |
|  | NO<sub>2</sub>/5 | / | / | / | / | / | 1.41(1.36, 1.45) * |
| Shanley et al. | PM<sub>10</sub>/11.1 | 1.43(1.21, 1.66) * | 2.42(1.09, 3.76) * | / | 1.18(0.81, 1.56) * | 0.18(-0.32, 0.68) * | / |
| Cai et al. | PM<sub>10</sub>/2 | / | 1.9(1.4, 2.4) * | / | / | 0.04(-0.2, 0.3) | / |
|  | NO<sub>2</sub>/7.4 | / | 2.1(1.6, 2.7) * | / | / | 0.4(0.1, 0.7) * | / |
| Mette et al. | PM<sub>2.5</sub>/1 | 0.70(0.12, 1.28) * | / | / | / | / | / |
|  | NO<sub>2</sub>/6.7 | 0.72(0.11, 1.34) * | / | / | / | / | / |

**Abbreviation:** TC indicates total cholesterol; TG, triglycerides; HTG, hypertriglyceridemia; LDL-C, low density lipoprotein; HDL-C, high density lipoprotein; MetS, metabolic syndrome.
\* means statistically significant association (p < 0.05).

**Table 4.**
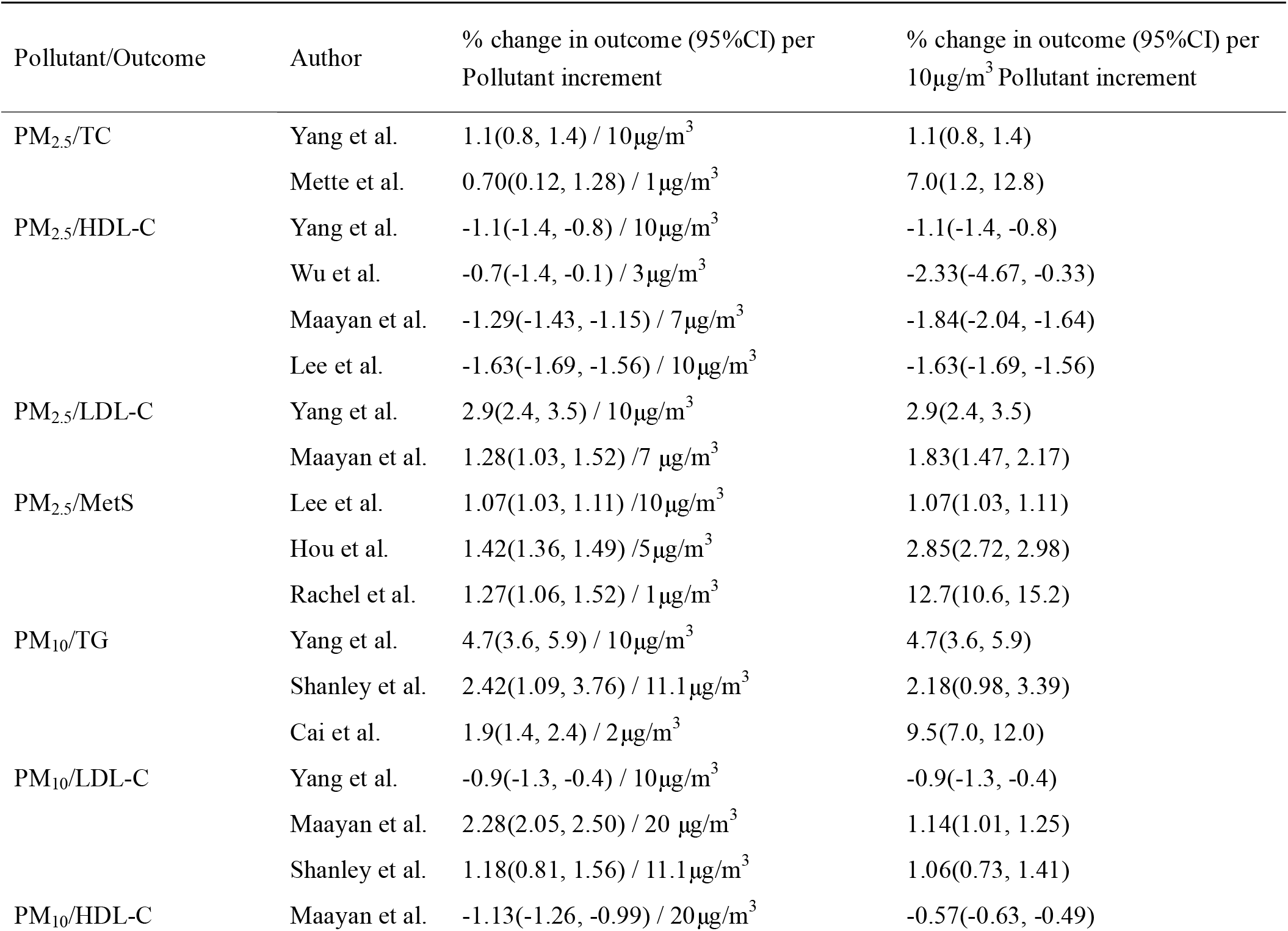
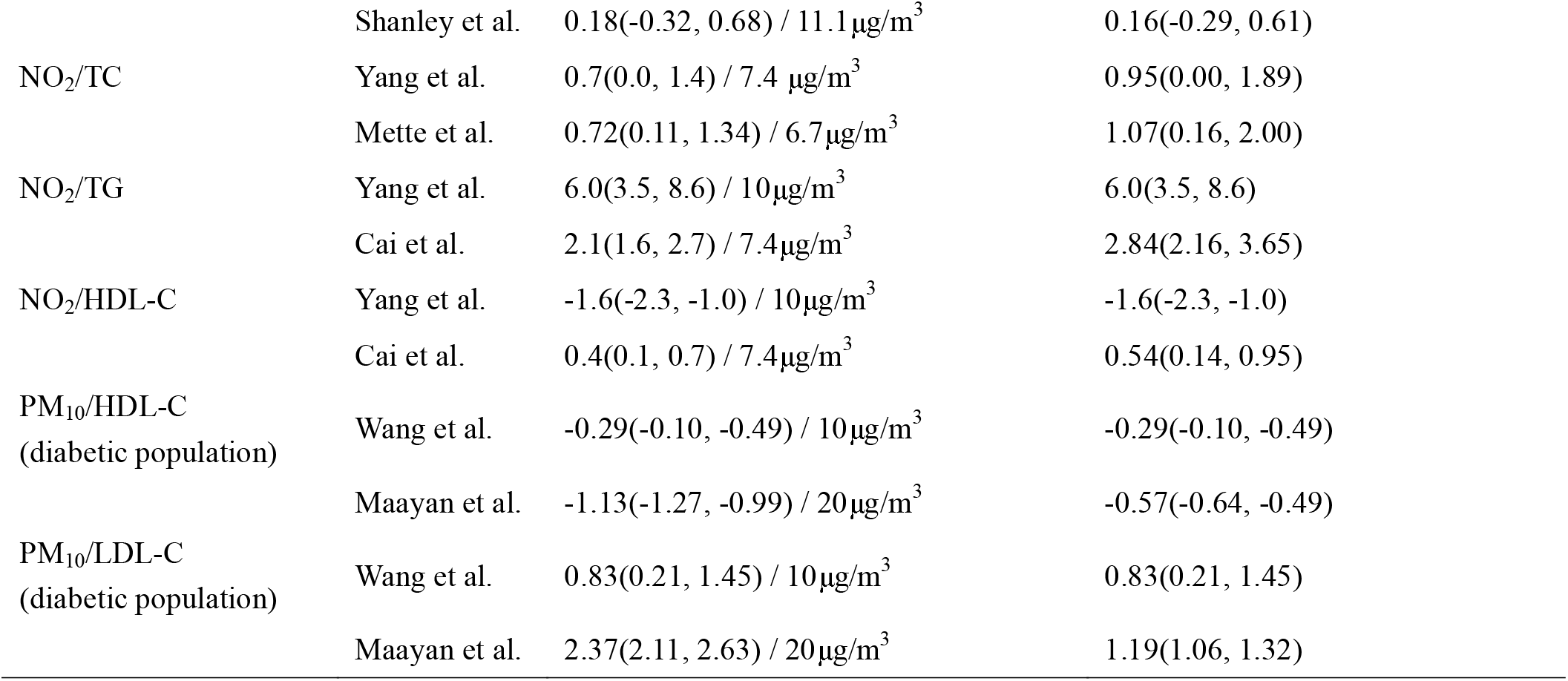
Data grouped by pollutants and data conversion.

### 3.4 Quantitative Meta-analysis

Based on a quantitative comparison of the ten studies included in the analysis, a total of twelve groups were identified to characterize the relationship between air pollutants and lipid parameters, including PM_2.5_/TC; PM_2.5_/HDL-C; PM_2.5_/LDL-C; PM_2.5_/MetS; PM_10_/TG; PM_10_/LDL-C; PM_10_/HDL-C; NO_2_/TC; NO_2_/TG; NO_2_/HDL-C; PM_10_/HDL-C (diabetic population); PM_10_/LDL-C(diabetic population).After data conversion, these effect estimates were analyzed using twelve different analyses for each pollutant/outcome group.

#### 3.4.1 PM_2.5_ with TC, HDL-C, LDL-C, and MetS^26, 27, 36, 39^

PM_2.5_ exposure was significantly associated with TC, HDL-C, LDL-C, and MetS. Specifically, for each 10 μg/m^3^ increase in PM_2.5_, TC levels increased by 3.31% (95% CI: −0.0229~0.0891, P=0.046), LDL-C levels increased by 2.34% (95% CI: 0.0130~0.0339, P=0.001), HDL-C levels decreased 1.57% (95% CI: −0.0185~-0.0128, P=0.001), and the MetS increased by 4.33% (95% CI: 0.0269~0.0598, P<0.001) (See Fig. 2).

**Fig.2.**
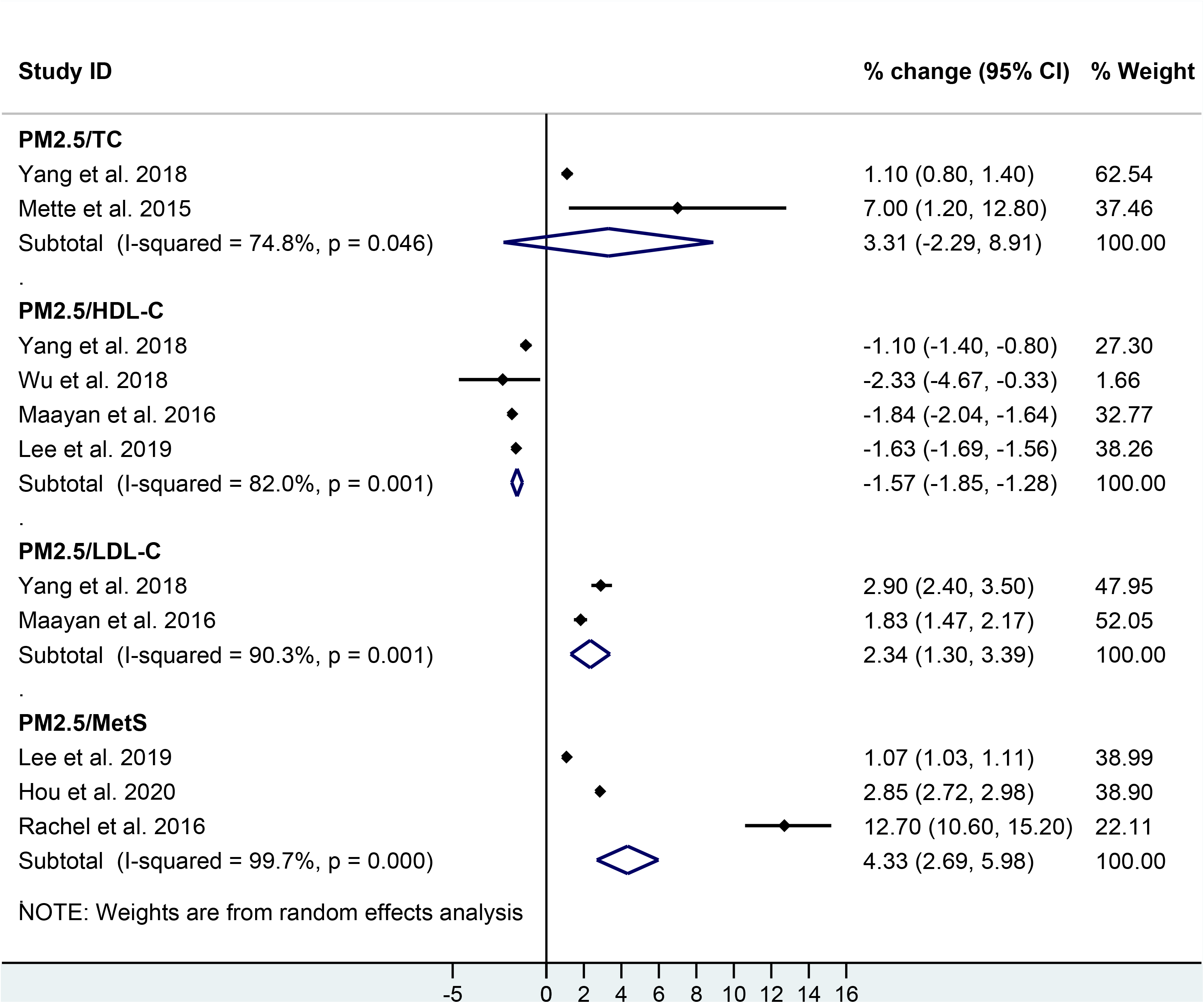
Forest plot of PM_2.5_ exposure (per 10 μg/m^3^ increase) with changes of TC, HDL-C, LDL levels and MetS morbidity.

#### 3.4.2 PM_10_ with TG, HDL-C, and LDL-C^26, 29, 35, 40^

PM_10_ exposure was significantly associated with TG, HDL-C, LDL-C. Specifically, for each 10 μg/m^3^ increase in PM_10_, TG levels increased by 5.27% (95% CI: 0.0203~0.0850, P<0.001), HDL-C levels decreased by 0.24% (95% CI: −0.0095~0.0047, P=0.002), and LDL-C levels increased by 0.45% (95% CI: −0.057~0.0147, P<0.001) (See Fig. 3).

**Fig.3.**
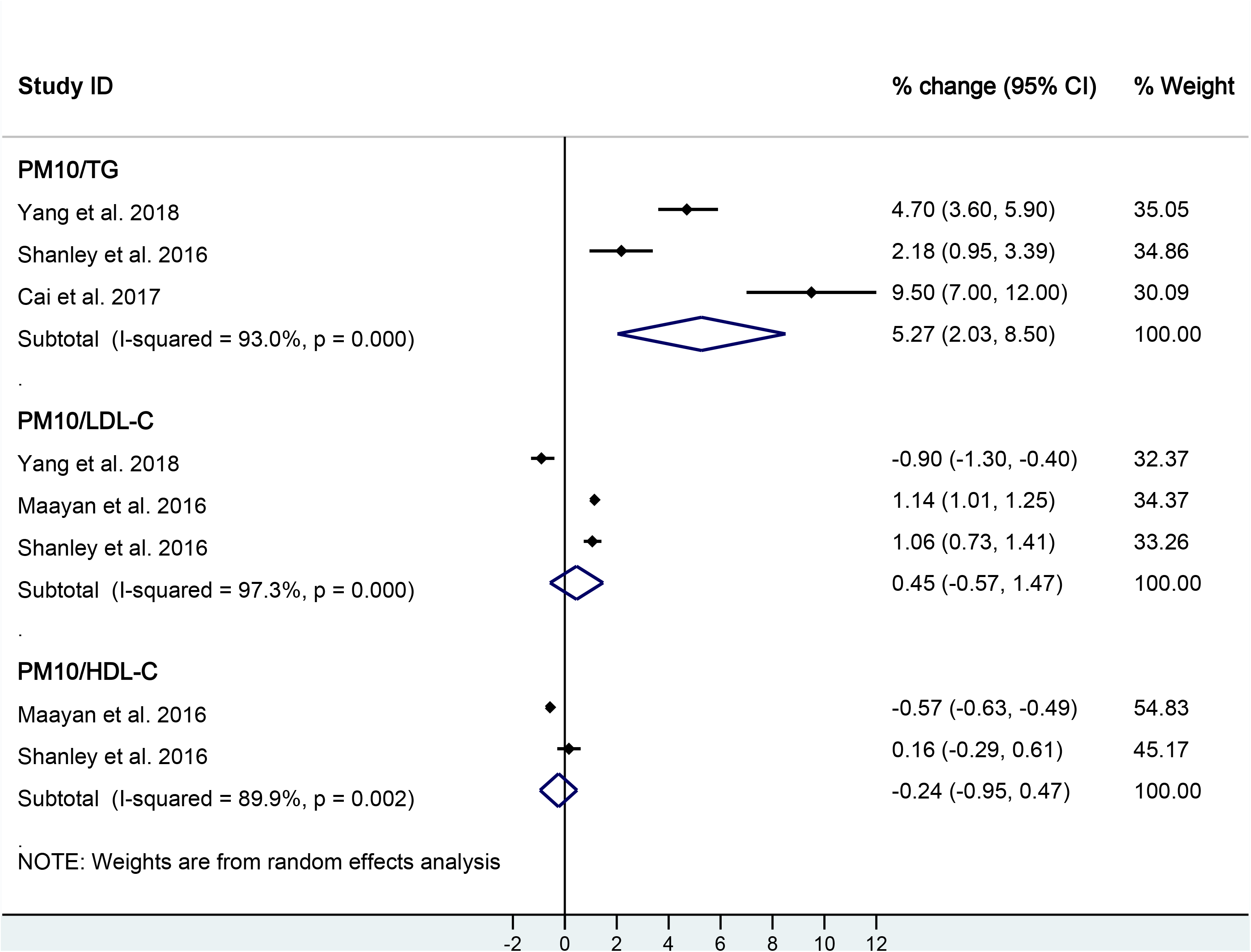
Forest plot of PM_10_ exposure (per 10 μg/m^3^ increase) with changes of TG, LDL-C, and HDL-C levels.

#### 3.4.3 NO_2_ with TG, HDL-C, and TC^26, 29, 39^

NO_2_ exposure was associated with TG and HDL-C; however, was not significantly associated with TC. Specifically, for each 10 μg/m^3^ increase in NO_2_, TG levels increased by 4.18% (95% CI: 0.0112~0.0723, P=0.020), and HDL-C levels decreased by 0.51% (95% CI: −0.0261~0.0158, P<0.001). TC levels increased by 1.01% (95% CI: 0.0035~0.0167, P=0.858) (See Fig. 4).

**Fig.4.**
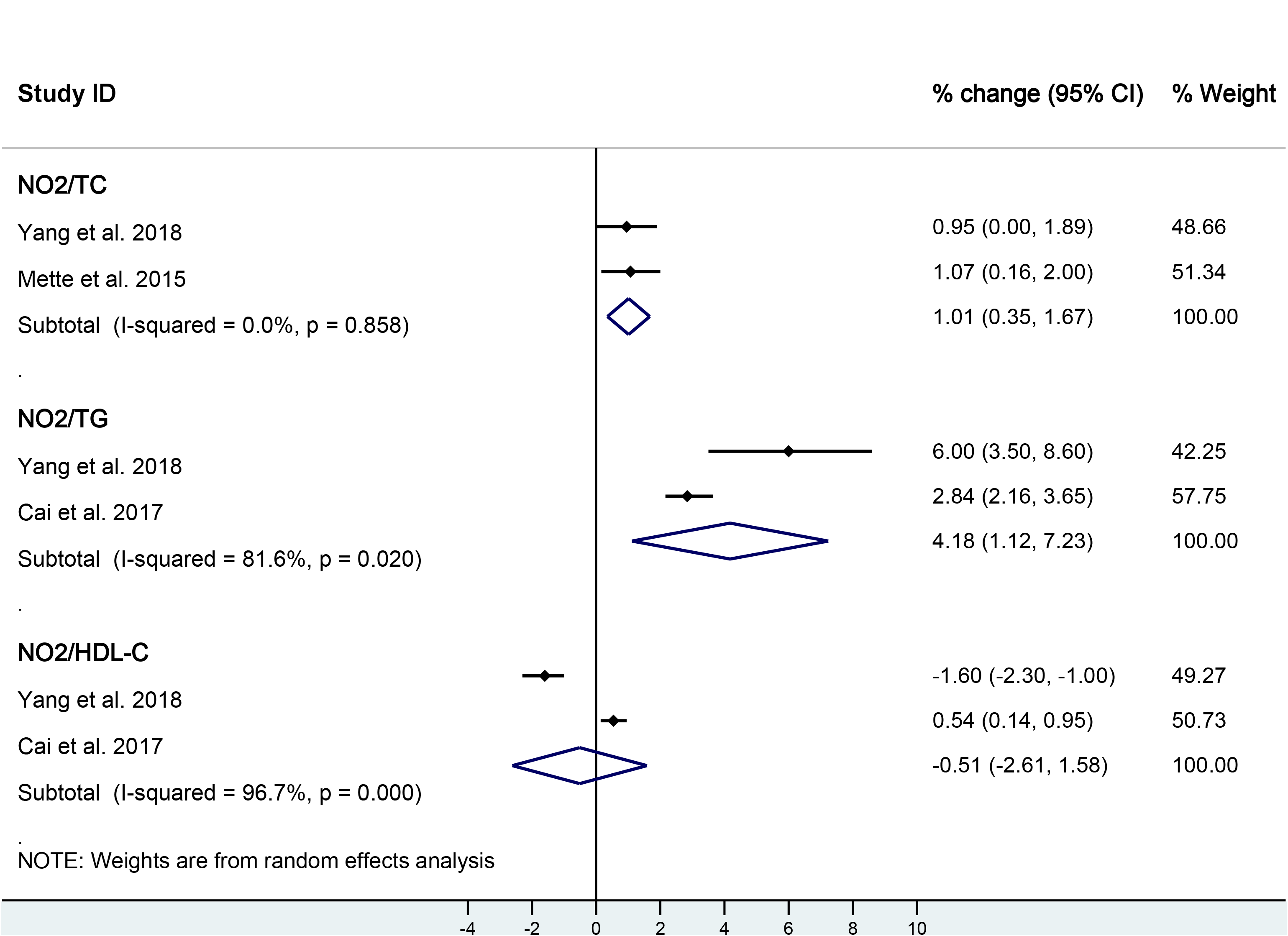
Forest plot of NO_2_ exposure (per 10 μg/m^3^ increase) with changes of TC, TG and HDL-C levels.

#### 3.4.4 PM_10_ with LDL-C and HDL-C in diabetic population^35, 37^

Three studies included subjects with diabetes mellitus, suggesting differential modification of LDL-C and HDL-C levels associated with PM_10_ exposure. PM_10_ exposure was significantly associated with HDL-C levels, but not with LDL-C levels. For each 10 μg/m^3^ increase in PM_10_, HDL-C levels decreased 0.45% (95% CI: −0.0072~-0.0017, P=0.009), while LDL-C increased by 1.14% (95% CI: 0.0090~0.00138, P=0.265) (See Fig. 5).

**Fig.5.**
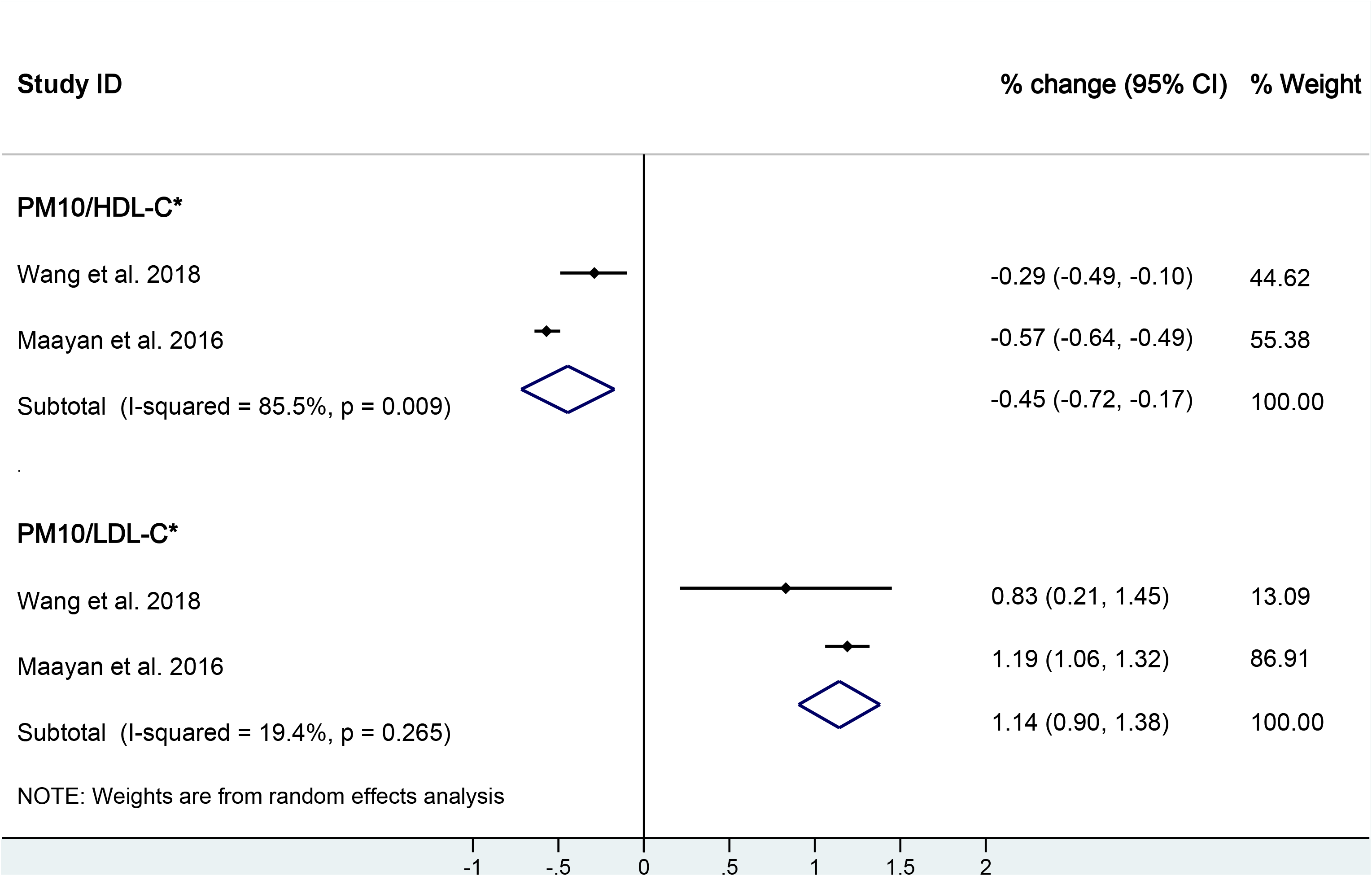
Forest plot of PM_10_ exposure (per 10 μg/m^3^ increase) with changes of HDL-C and LDL-C levels in diabetic population.

## 4 Discussion

Significant associations were acquired in this study between air pollutants and lipoprotein parameters, although some with evidence of substantial between-study heterogeneity. Particularly in subgroup of PM_2.5_/MetS, the result may add to evidence that MetS may increase susceptibility to cardiovascular morbidity associated with air pollution exposure.

It’s reported ^25^ that LDL-C and TG levels increased by 0.12% (95% CI: 0.0178~0.0206) and 3.14% (95% CI: 0.0136~0.0495) respectively, per 10 μg/m^3^ increase in PM_10_, which is consistent with our study, although effect estimates were smaller than the current study. They also observed a decrease of 0.45% (95% CI: −0.0265~-0.0181) in HDL-C levels per 10 μg/m^3^ increase in NO_2_, but no statistically significant changes in TC. When considering the differences between the two studies, such factors as the use of differential inclusion and exclusion criteria for screening may have resulted in more or less robust records for final selection. To the extent that studies focused on associations between air pollutants and lipometabolic disturbance were much less based on the pre-search, the inclusion criteria in our study were broader, while the exclusion criteria were similar to the meta-analysis by Vaio et al.. As a result, our study included an additional study ^35^ in the subgroup category for studies investigating the relationship between PM_10_ exposures and LDL-C and TG levels. In this study, PM_2.5_ was more significantly associated with multiple lipid parameters compared to PM_10_, which is consistent with findings by Hazrije et al.^41^; however, they reported a stronger association between myocardial infarction and PM_2.5_ compared to PM_10_. This may result from the differential characteristics of PM_2.5_ such as smaller diameter size, larger surface area, slower sedimentation rate, and longer retention time in the air compared to PM_10_.

This paper reported that elevated PM_10_ exposure on the diabetic population was associated with higher HDL-C levels, while not significantly associated with LDL-C levels. This was only partially in accordance with the previous study^42^, which reported significant associations between PM_10_ exposure and both HDL-C levels and LDL-C levels. Such discrepancy may be due to differences in the characteristics of the study population. Further, two studies^35, 37^ included in this meta-analysis contained subjects with diabetes using data obtained from medical institutions, whereas Wang et al.^42^ had the advantage of follow-up throughout the study period, which may have provided a more precise measure of changes in lipid parameters.

The present studies suggest that air pollution may adversely affect lipometabolic balance by promoting LDL oxidation, disrupting the scavenger receptor and LDL receptor body function, or accelerating the accumulation of lipids in plaque^43, 44^. HDL, as a protective factor for lipid metabolic disorders, decreased in levels when exposed to air pollutants^45^.

Despite a considerably inclusive search strategy, with few restrictions on study design and population characteristics, our final selection yielded 10 qualified studies for this meta-analysis. We found high between-study heterogeneity in most of the subgroups studied, which, taken together, may attenuate credibility with cumulative evidence. Yet, heterogeneity may be captured by unmeasured variables in some studies that were not reported such as population susceptibility, temperature, lipid-lowering medications, distribution of air pollutants or other potential sources^46, 47^.

There are some limitations that should be addressed. First, most of the literature failed to consider whether subjects had taken lipid-lowering drugs or related drugs using history before or during the study period, which may bias results and lead to less positive change in blood lipid parameter levels^48^. Furthermore, evidence suggests that noise intensity can directly impact lipid parameters levels^29, 39, 49-52^. Considering noise has obvious geographical distribution characteristics, i.e., urban residential areas typically have higher levels of noise intensity than rural or remote areas, and that distance between residential areas and main roadways could fluctuate multiple lipid parameters, future analyses should further examine noise intensity as a possible confounding risk factor. Similarly, green spaces near residential areas or workplaces have been shown to influence lipometabolic balance and should additionally be included as a confounding variable^52-54^. Moreover, residential greenness has been shown elsewhere to have a beneficial effect on MetS and diabetes; accordingly, this association could be attenuated after adjustment for air pollution^55, 56^. To quantify air pollution exposure, many studies^27, 29, 37, 39, 40^ use ground-based measurements derived from fixed air quality monitoring sites. While other studies^26, 35, 36, 38^ incorporate predictive models estimated from spatiotemporal hybrid modeling. More specifically, models may utilize low-cost GPS on mobile devices to monitor an average individual exposure to air pollutants in real time, over the study period. Such methods greatly increase the accuracy of measurements and may reflect a better estimate of true exposure to air pollutants. Despite the limitations, previous studies inclined to investigate the relationship between blood lipid parameters levels and a single air pollutant. Examining the effects of air pollution exposure one pollutant at a time underestimates the complexity of atmospheric chemical mixing and the multiple pollutant sources implicated in the link between air pollution exposure and adverse health, and more specifically, the nuanced effects of various pollutants on blood lipid parameters.

## 5 Conclusion

This study found that PM_10_, PM_2.5_, and NO_2_ were significantly associated with HDL-C, LDL-C, TC, and TG levels, suggesting a link between air pollution and cardiovascular morbidity. Therefore, improving air quality may yield substantial health benefits.

## Data Availability

This study was conducted using standard methods which follow the Preferred Reporting Items for Systematic Review and Meta-Analysis (PISMA)(J et al., 2020; "Preferred reporting items for systematic review and meta-analysis protocols (PRISMA-P) 2015: elaboration and explanation," 2016). Seven electronic databases, including China National Knowledge Infrastructure (CNKI), Wanfang, Vip, SinoMed, Pubmed, EMBASE, and the Cochrane Library, were searched for peer-reviewed articles published from 2000 to January 28, 2020. Keywords searched included: ("Air Pollution" OR "Pollution, Air" OR "Air Quality" OR "Ultrafine Fibers" OR "Airborne Particulate Matter" OR "Particulate Matter, Airborne" OR "Air Pollutants, Particulate" OR "Particulate Air Pollutants" OR "Ambient Particulate Matter" OR "Particulate Matter, Ambient" OR "Ultrafine Particulate Matter" OR “Particulate Matter, Ultrafine” OR "Ultrafine Particles" OR "Particles, Ultrafine") AND ("Cardiovascular Disease" OR "Disease, Cardiovascular" OR "Diseases, Cardiovascular" OR "HDL" OR "LDL" OR "cholesterol" OR " cholesterin" OR "cholesteric" OR "TC" OR "TG" OR "dyslipidaemia" OR "HDL-C" OR "LDL-C") AND ("Cohort" OR "Cross-sectional" OR "Case control" OR "Case-control" OR "Epidemiology OR "Epidemiological"). Additionally, references of included literature and one previously published systematic review were manually retrieved.

https://pubmed.ncbi.nlm.nih.gov/27218271/

https://doi.org/10.1016/j.envint.2018.07.016

https://doi.org/10.1016/j.scitotenv.2018.11.149

https://doi.org/10.3390/ijerph15040631

https://doi.org/10.1093/aje/kww157

https://doi.org/10.1016/j.envint.2015.09.021

https://www.ncbi.nlm.nih.gov/pubmed/20853625

https://doi.org/10.1097/ede.0000000000000426

https://doi.org/10.1016/j.ijheh.2019.01.010

https://doi.org/10.1016/j.envint.2020.105459

## Acknowledgment

This work was supported by the Scientific Research Project of Beijing Educational Committee (KM201910025023), Open Foundation of Beijing Key Laboratory of Occupational Safety and Health (2019KJ000377), Beijing Academy of Science and Technology - Reform and Development (PY2020HJ34), and HERCULES Exposome Research Centre (P30ES019776).

---

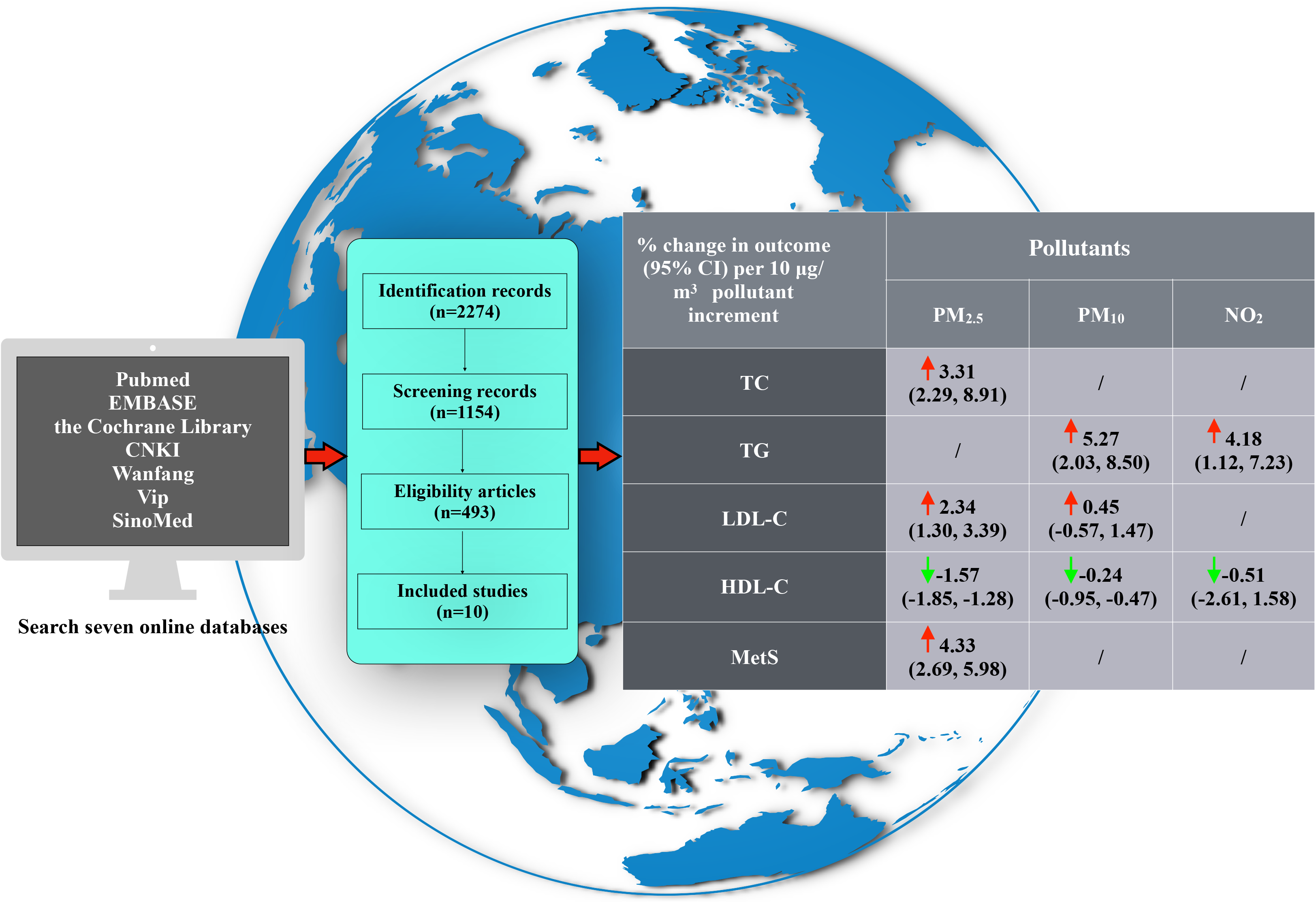

